# A homozygous *ATP2A2* variant alters sarcoendoplasmic reticulum Ca^2+^-ATPase 2 function in skeletal muscle and causes a novel vacuolar myopathy

**DOI:** 10.1101/2024.05.14.24307284

**Authors:** Laura Llansó, Gianina Ravenscroft, Cristina Aceituno, Antonio Gutiérrez, Jevin Parmar, Pia Gallano, Marta Caballero-Ávila, Álvaro Carbayo, Ana Vesperinas, Roger Collet, Rosa Blanco, Nigel Laing, Leif Hove-Madsen, Eduard Gallardo, Montse Olivé

## Abstract

**Background:** Sarcoendoplasmic reticulum Ca^2+^-ATPase isoform 2 (SERCA2), encoded by *ATP2A2*, is a key protein involved in intracellular Ca^2+^ homeostasis. The transcript SERCA2a is predominantly expressed in cardiac muscle and in type I myofibers, while SERCA2b is ubiquitously expressed including in skin cells. To date, variants in this gene were reported to be the cause of Darier disease, an autosomal dominant dermatologic disorder, but have never been linked to primary skeletal muscle disease. We describe four patients suffering from a novel hereditary myopathy caused by a homozygous missense variant in *ATP2A2*.

**Methods:** We studied a family with four affected individuals suffering from an adult-onset progressive skeletal myopathy. We performed a comprehensive evaluation of the clinical phenotype, serum CK levels, muscle MRI, and muscle biopsy, with genetic workup by means of gene panel sequencing followed by whole genome sequencing and segregation analysis. Immunohistochemistry and western blot (WB) to evaluate SERCA2 and SERCA1 expression in skeletal muscle was performed. We evaluated kinetics of Ca^2+^handling following caffeine exposure or voltage-induced sarcolemma depolarization in patient myoblasts and myotubes, compared to healthy controls.

**Results:** Four siblings in their fifties developed in early adulthood symmetric proximal weakness in lower limbs, which was slowly progressive over time. They had no skin or cardiac involvement. Biopsy findings in two affected individuals showed small vacuoles restricted to type I myofibers. Ultrastructural analysis showed dilation and proliferation of T-tubules, swelling of sarcoplasmic reticulum and autophagic vacuoles. Genome sequencing revealed a homozygous variant in *ATP2A2* (c.1117G>A, p.(Glu373Lys)) which segregated with the disease. Immunohistochemistry suggested SERCA2 mislocalization in patient myofibers compared to controls. WB did not show changes in the amount or molecular weight of the protein. *In vitro* functional studies revealed delayed sarcoendoplasmic reticulum Ca^2+^reuptake in patient myotubes, consistent with an altered pumping capacity of SERCA2 after cell stimulation with caffeine or depolarization.

**Conclusions:** We report a novel adult-onset vacuolar myopathy caused by a homozygous variant in *ATP2A2*, resulting in a pure skeletal muscle phenotype with a limb-girdle distribution. Biopsy findings and functional studies demonstrating an impaired function of SERCA2 and consequent Ca^2+^ dysregulation in slow-twitch skeletal myofibers highly support the pathogenicity of the variant.

## Introduction

Sarcoendoplasmic reticulum Ca^2+^-ATPases (SERCA) are Ca^2+^ pumps located in the sarcoendoplasmic reticulum (SR) membrane. SERCA pumps are encoded by three genes in humans (*ATP2A1, ATP2A2, ATP2A3*) that generate multiple mRNA splice variants and 10 protein isoforms (SERCA1a,b; SERCA2a-c; SECA3a-f)(1–4). Together with the highly related SERCA1 and SERCA3 isoforms encoded by *ATP2A1* and *ATP2A3*, respectively, SERCA2 belongs to the large family of P-type cation pumps that couple ATP hydrolysis with active Ca^2+^ transport across membranes. SERCA channels are 110-kDa transmembrane proteins that specifically maintain low cytosolic Ca^2+^ concentrations by actively transporting Ca^2+^ from the cytosol into the SR, having an important role in excitation-contraction coupling in muscle. Thus, they carry out a dual function by lowering cytosolic Ca^2+^ concentrations to allow muscle relaxation, and, by restoring Ca^2+^ stores in the SR, they allow subsequent myofiber contraction. The opposite function of Ca^2+^ release to the cytosol is performed by ryanodine receptors (RYR1 in skeletal muscle), also located in the SR membrane. The *ATP2A1-3* gene family is highly conserved among species, as SERCA pumps are present in all living organisms including plants, yeast, invertebrates and mammals(5). Although the main function of SERCA is to control cytosolic Ca^2+^, it also plays a vital role in other cellular functions including cell growth and differentiation in invertebrates, plants and yeast(6–8). The main SERCA regulatory proteins are sarcolipin and phospholamban(9).

SERCA isoform levels depend on tissue-specific expression and alternative splicing; abundance of these isoforms also changes during development and in normal aging (1–4). For instance, in human adults, SERCA1a is predominantly present in fast-twitch (type II) skeletal myofibers, while SERCA2a is found mostly in slow-twitch (type I) skeletal myofibers and myocardial tissue. SERCA2b is ubiquitously expressed in all tissues at low levels, including smooth muscle and skin, and SERCA3 isoforms are expressed in a variety of non-muscle cells(5,10). In addition, numerous studies of animal skeletal muscle have shown that extrinsic factors such as induced denervation or chronic low-frequency electrical stimulation influence the expression pattern of SERCA isoforms(11–14). The ubiquitous SERCA2b isoform is the SERCA pump with the highest Ca^2+^ affinity but with a lower turnover rate of the pump, due to a 2b-tail that stabilizes the E1 structural conformation of the protein with high-affinity Ca^2+^-binding sites facing the cytoplasm(15).

To date, over 270 heterozygous pathogenic *ATP2A2* variants disrupting critical functional domains of SERCA2 have been associated with Darier disease (DD, OMIM 124200), a skin condition also called keratosis follicularis(16,17). DD is an autosomal dominant disease characterized by dysfunction in keratinocyte adhesion leading to hyperkeratotic papules in seborrheic areas, palmoplantar pits, and distinctive nail abnormalities. Ca^2+^ dysregulation, endoplasmic reticulum stress and abnormal protein folding leading to cellular apoptosis and acantholysis in keratinocytes are the main mechanisms described in DD pathophysiology(18–22).

On the other hand, biallelic variants in the *ATP2A1* gene cause Brody disease (OMIM 601003), a rare autosomal recessive myopathy characterized by exercise-induced muscle stiffness. Patients with Brody disease frequently have an athletic appearance and suffer from muscle cramps and generalized myalgia, with worsening of symptoms upon exposure to cold temperatures. Nevertheless, they rarely develop muscle wasting or weakness(23–28).

While *ATP2A1* biallelic variants are linked to Brody disease, mutations in *ATP2A2* have never been reported to be causative of muscle disease, and DD is not associated with muscle weakness.

We herein report a family with four affected siblings suffering from progressive proximal muscle weakness associated with a homozygous missense pathogenic variant located in exon 9 of *ATP2A2*. We describe the clinical and myopathological phenotype and provide *in vitro* functional studies supporting the pathogenicity of the identified variant.

## Materials and methods

### Patients

We have clinically characterized four affected siblings from a single family (II:1, II:2, II:3 and II:4). Patients presented with an autosomal recessive slowly progressive proximal myopathy. Symptom onset was variable between the third and fourth decades. We performed pedigree analysis, neurological examination, including muscle strength evaluation according to Medical Research Council (MRC) grading scale, cardiac evaluation comprising EKG and cardiac ultrasound, serum CK levels from all affected individuals and EMG in individuals II:1 and II:3.

The study was approved by the ethics committees of the participating institutions. Sample collection was performed after written informed consent from the patients according to the declaration of Helsinki.

### Muscle biopsy

Muscle biopsy was performed in two affected individuals (II:1 and II:4). The samples were immediately frozen in liquid cooled isopentane. Cryostat sections were processed for routine histological and histochemical techniques and for cholinesterase activity. In addition, muscle sections from individual II:1 were processed for SERCA2, SERCA1, RYR1, p62, caveolin-3, dystrophin, and dysferlin immunohistochemistry or immunofluorescence using standard procedures. In addition, SERCA2 western blot (WB) was performed in sample from individual II:1 as previously described(29). A summary of the antibodies used in the present study is provided in Supplementary table T1. Finally, a small sample of muscle from individual II:1 was fixed in 2% glutaraldehyde, postfixed with 1% osmium tetroxide, and embedded in araldite. Ultrathin sections were stained with uranyl acetate and lead citrate and viewed with a JEOL 1011 electron microscope.

### Muscle MRI

Whole-body muscle magnetic resonance imaging (MRI) was performed in 3 patients. The MRI protocol included T1-weighted (T1w), short-T1 inversion recovery (STIR), and T2-weighted (T2w) sequences.

### Molecular genetics

The proband was initially tested for a panel that includes 137 genes associated with muscular dystrophies, congenital myopathies and congenital myasthenic syndromes with negative results. Subsequent clinical exome sequencing detected no candidate variants.

Whole genome sequencing (WGS) was performed at the Kinghorn Centre for Clinical Genetics, on genomic DNA isolated from all four affected members, the unaffected sibling, and the unaffected parents. The Illumina DNA PCR-free library preparation kit is what was used for the library preparation and sequencing was performed using a S4 300 cycle flow cell on the NovaSeq 6000 sequencer.

WGS data processing was performed by the Centre for Population Genomics following the DRAGEN GATK best practices pipeline. Reads were aligned to the hg38 reference genome using Dragmap (v1.3.0). Cohort-wide joint calling of single nucleotide variants (SNVs) and small insertion/deletion (indel) variants was performed using GATK HaplotypeCaller (v4.2.6.1) with “—dragen-mode” enabled. Variants were annotated using VEP 105, and loaded into the web-based variant filtration platform, Seqr(30). Sample sex and relatedness quality checks were performed using Somalier (v0.2.15)(31).

Within Seqr, variant curation was performed, running a filter for all recessive, rare (allele frequency <0.001 in gnomAD and TOPMed) coding and likely coding changes (including extended splice region variants) with high call quality (genotype quality 20 and allele balance of 0.25).

### Multiple sequence alignment

To examine the conservation of the p.Glu373Lys substitution, multiple sequence alignment of SERCA2 orthologs was performed. SERCA2 protein sequences for each species were retrieved from the UniProt database (https://www.uniprot.org/) using their respective accession codes (Supplementary table T2). Alignments were performed using the default MAFFT algorithm in Jalview v2.11.3.2 (https://www.jalview.org/).

### Protein structure modelling and *in silico* mutagenesis

The experimentally determined protein structure of SERCA2 (32) was retrieved from the RCSB Protein Data Bank, Accession 5ZTF (https://www.rcsb.org/). The three-dimensional structure was visualised using PyMol v2.5.2 (https://pymol.org/2/). To investigate variant effect on three-dimensional protein structure, in silico mutagenesis for the identified missense substitution was performed through the PyMol “mutagenesis” function.

### Functional studies to assess Ca^2+^ kinetics after sarcolemma depolarization in myoblasts and myotubes

For cell culture studies, primary myoblasts were isolated, cultured and differentiated as previously described(33). To measure cytoplasmic Ca^2+^, cell cultures were incubated with 2 µM Fluo-4 (Invitrogen) for 20 minutes at RT, followed by wash (twice) with a physiological buffer containing 132 mM NaCl, 0,33 mM NaH2PO4, 4 mM KCl, 4 mM NaHCO3, 2 mM CaCl2, 1,6 mM MgCl2, 10 mM HEPES, 5 mM glucose and 5 mM pyruvic acid. pH was adjusted to 7.4 with NaOH. Experiments were performed at room temperature. Images (256×256 pixels) were recorded with a resonance scanning confocal microscope Leica SP5 AOBS (Wetzlar, Germany) and a HCX PL APO CS 20.0×0.70 IMM objective at a frame rate of 52 images/s.

Electrical field stimulation (15 V, 2ms square pulses) was used to elicit Ca^2+^ transients using a Hugo Sachs Elektronik stimulator type 223 (Harvard Apparatus). The stimulation frequency was increased stepwise by reducing the stimulation interval from 20s to 10s, 3s, and 1s. To measure Ca^2+^ release from the SR, cells were exposed transiently to 10 mM caffeine for 9 seconds. A custom-made algorithm was used to detect myotubes that responded to field stimulation or caffeine exposure, and to determine the baseline, amplitude, time to peak, duration and decay of the Ca^2+^ signals as previously described(34).

## Results

### Clinical phenotype

The proband is a woman in her fifties (II:1) from a small town. She was born from healthy, unrelated parents. Symptom onset was between 30-35 years old, when she first noticed difficulty in doing squats. She was previously a sporty person and she did not have other medical issues. Pelvic weakness slowly progressed over time. Twenty years after the initial symptoms, she started having some difficulties in raising up her arms. Currently, she has difficulties climbing stairs, and she is unable to run or jump. Despite her motor difficulties, she continues to perform exercise 5 times per week. She has three other siblings, twins in the sixth decade of life, and another in the fifth decade, with similar symptoms, and a younger asymptomatic sibling in the fourth decade of life (Fig.1: not exposed for Medrxiv requirements). The twin siblings, despite being younger than the proband, are more severely affected. Onset of weakness in all individuals ranged from 23 to 40 years. None of them had facial, bulbar or respiratory weakness. None had skin lesions. None had previous episodes of malignant hyperthermia, nor muscle stiffness. Cardiac workup including EKG and cardiac ultrasound did not show any abnormality. Physical examination of the affected relatives revealed mild scapular winging and variable degrees of limb-girdle weakness, more pronounced in lower limbs, with milder distal weakness affecting anterolateral leg compartments and finger extensors of forearms. They also presented mild ankle contractures and calf atrophy. No axial weakness was detected.

**Figure 1.** Family pedigree with *ATP2A2* mutations causing a proximal vacuolar myopathy. Black symbols represent affected members carrying the variant p.(Glu373Lys) in homozygosis. The black and white symbols represent unaffected members who were carriers of the variant in heterozygosis.

### Laboratory and electrodiagnostic studies

CK levels in the proband (II:1) were 900 IU/L and in the other three affected individuals were 533 IU/L (II:2), 900 IU/L (II:3) and 537 IU/L (II:4). Nerve conduction studies in patients II:1 and II:3 were normal. Needle electromyography showed myopathic changes with abundant spontaneous activity at rest in upper and lower limbs, as well as in paraspinal muscles. A dry blood spot screening for Pompe disease was negative.

### Muscle pathology and western blotting

Muscle biopsy of individuals II:1 and II:4 showed increased variability in fiber size, increased number of internal nuclei and abundant small vacuoles exclusively located in type I (slow-twitch) muscle fibers. Some of the vacuoles were limited by a membrane as revealed with anticholinesterase activity and with dystrophin, dysferlin, and caveolin-3 immunohistochemistry, suggesting they originated from the T-tubules. The sites occupied by the vacuoles were devoid of oxidative enzyme activity. ATPase activity revealed a mild type II myofiber predominance in both individuals. Muscle biopsy findings are shown in Fig.2.

**Figure 2.**
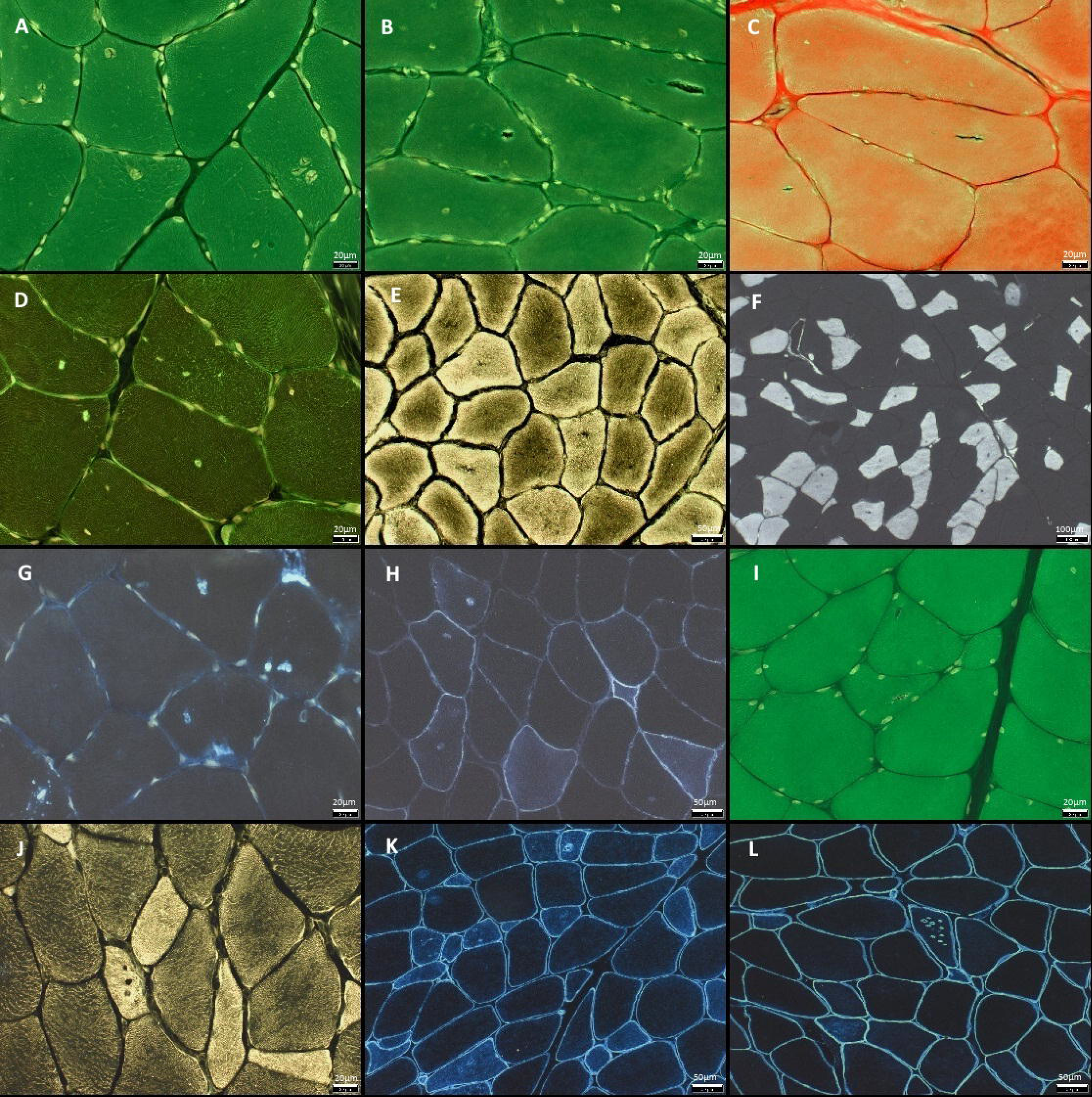
Muscle biopsy findings. A-H: images from patient II:1 (proband). I-L: images from patient II:4. **(A,B,I)** Hematoxylin*-*eosin stain shows increased variability in fiber size, increased number of internal nuclei, and single or multiple vacuoles exclusively located in type I myofibers. Some vacuoles appear optically empty and others filled by granular basophilic material. **(C)** The vacuoles have a reddish appearance on modified Gomori stain. **(D)** PAS stain shows positivity inside some of the vacuoles suggesting presence of glycogen. **(E,J)** Oxidative enzyme activity is absent in the areas occupied by the vacuoles as revealed with NADH reaction. **(F)** ATPase at pH 4.35 showing location of the vacuoles exclusively seen in type I (slow-twitch) fibers, and mild predominance of type II fibers. **(G)** p62 immunoreactivity is observed at the site of some vacuoles. **(H)** Cholinesterase activity is increased at the site of vacuoles. **(K)** caveolin-3 and **(L)** dystrophin immunohistochemistry show some of the vacuoles limited by a membrane. Moreover, caveolin-3 is overexpressed at the cytoplasm of some fibers.

Ultrastructural analysis in sample from individual II:1 revealed proliferation and dilatation of T-tubules, some of them leading to membrane-limited vacuoles containing membrane debris. There was sarcoplasmic reticulum proliferation with honeycomb-like structures and some autophagic vacuoles (Fig.3). Immunohistochemistry to assess SERCA2 expression showed a redistribution of SERCA2 with less protein expression in the cytoplasm but increased immunoreactivity at the sarcolemma and at the site of the vacuoles (Fig.4A,B). Western blot for SERCA2 did not show differences regarding the amount or molecular weight of the protein in the patient compared to a healthy control (Fig.4E). SERCA1 expression in the same sample was no different from a control muscle, except for a predominance of SERCA1-positive myofibers which corresponds to the type II myofiber predominance observed in the ATPase staining (Fig.4C,D). RYR1 immunofluorescence did not reveal different expression in patient muscle compared to control muscle (data not shown).

**Figure 3.**
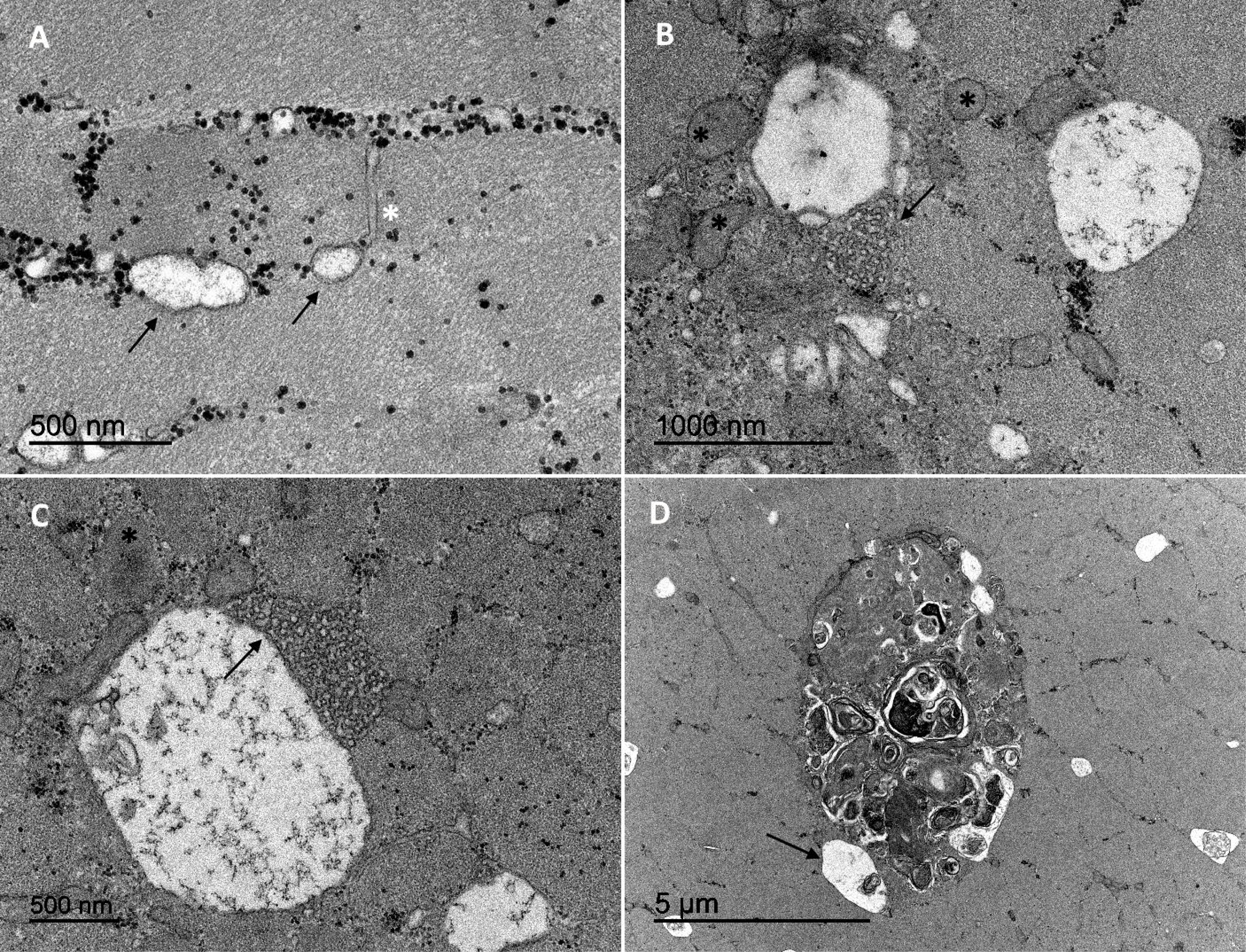
Electron microscopy images. Ultrastructural analysis of sample from individual II:1. Image **(A)** shows a T tubule (asterisk) next to membrane-bound vacuoles (arrows) and numerous glycogen granules. **(B)** Two large vacuoles, in combination with distended sarcoplasmic reticulum (asterisks), and honeycomb-like structures (arrow) corresponding to proliferation of the tubular system. Similar findings are shown in image **(C)** at higher magnification. **(D)** A large autophagic vacuole containing myelin figures and residual bodies. Some clear vacuoles are seen at the periphery (arrow).

**Figure 4.**
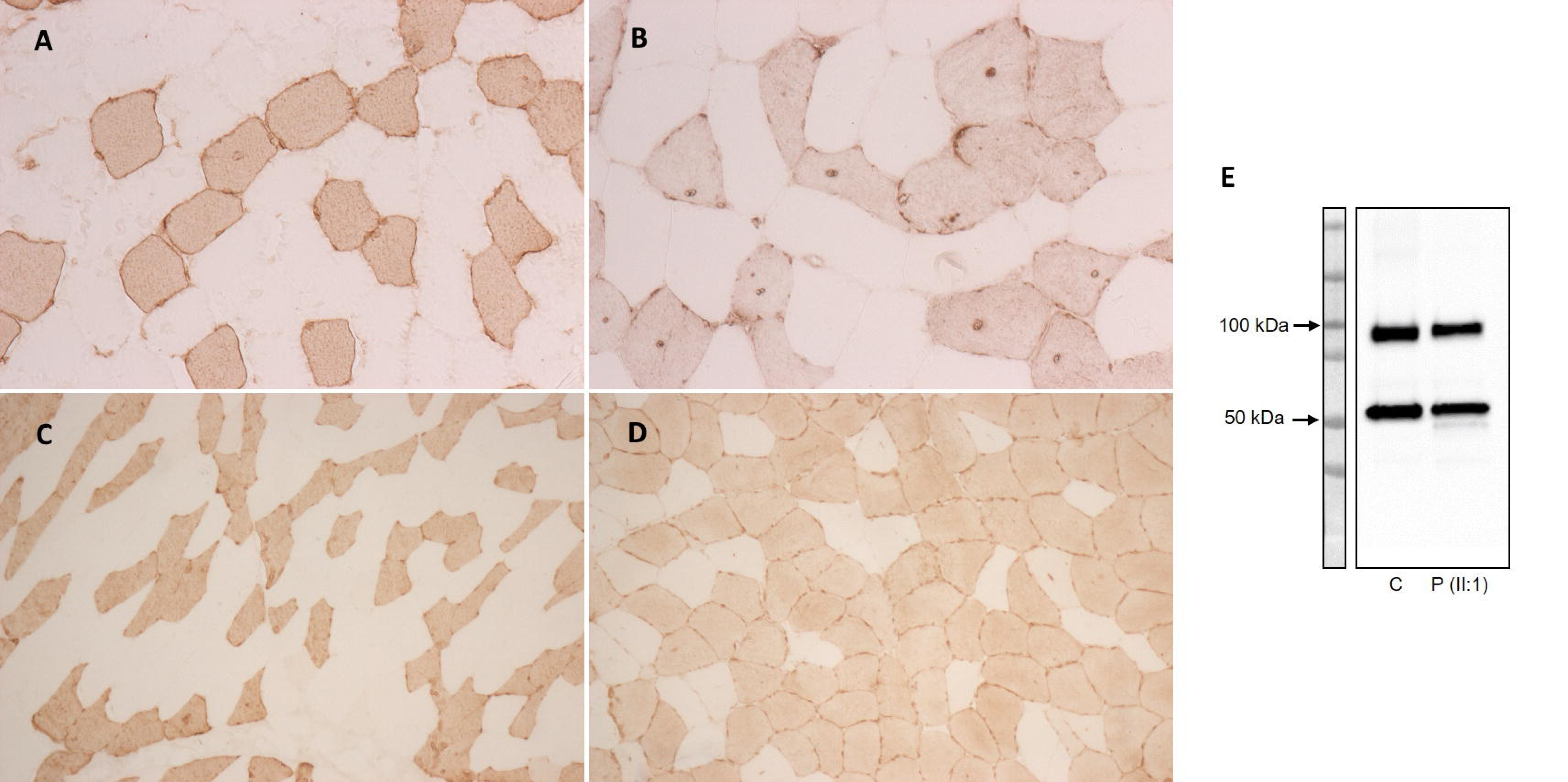
SERCA2 and SERCA1 expression in patient muscle compared to a healthy control. Immunohistochemical analysis for SERCA2 in control muscle **(A)** and patient II:1 muscle **(B)** reveals a less intense immunoreactivity at the cytoplasm and increased immunoreactivity at the site of the vacuoles and in some subsarcolemmal areas in patient type I myofibers, suggesting a redistribution of the protein. Immunohistochemistry for SERCA1 in control muscle **(C)** and patient muscle **(D)** shows a similar mosaic pattern of expression representing type II muscle fibers. Note the predominance of SERCA1-positive fibers in the patient, corresponding to type II fiber predominance. **(E)** Western blotting for SERCA2 (∼110 kDa) does not show differences regarding the total amount of protein or molecular weight, in patient muscle compared to the control. Desmin (∼54 kDa) was used as a protein loading control. C: control. P: patient. Used antibodies and dilutions are described in Supplementary figure 1.

### Muscle MRI

Muscle MRI showed variable degrees of atrophy and fat replacement in pelvic and thigh muscles with a notably sparing and hypertrophy of adductor longus and rectus femoris in all patients. In the leg compartments, calf muscles were more affected than the anterior compartment muscles. Mild involvement of scapular girdle was present in all siblings, mainly affecting deltoids, infraspinatus, and subscapularis muscles. Thoracic and lumbar paravertebral muscles were also moderately involved. Whole-body T1w muscle images from three affected individuals are shown in Fig.5.

**Figure 5.**
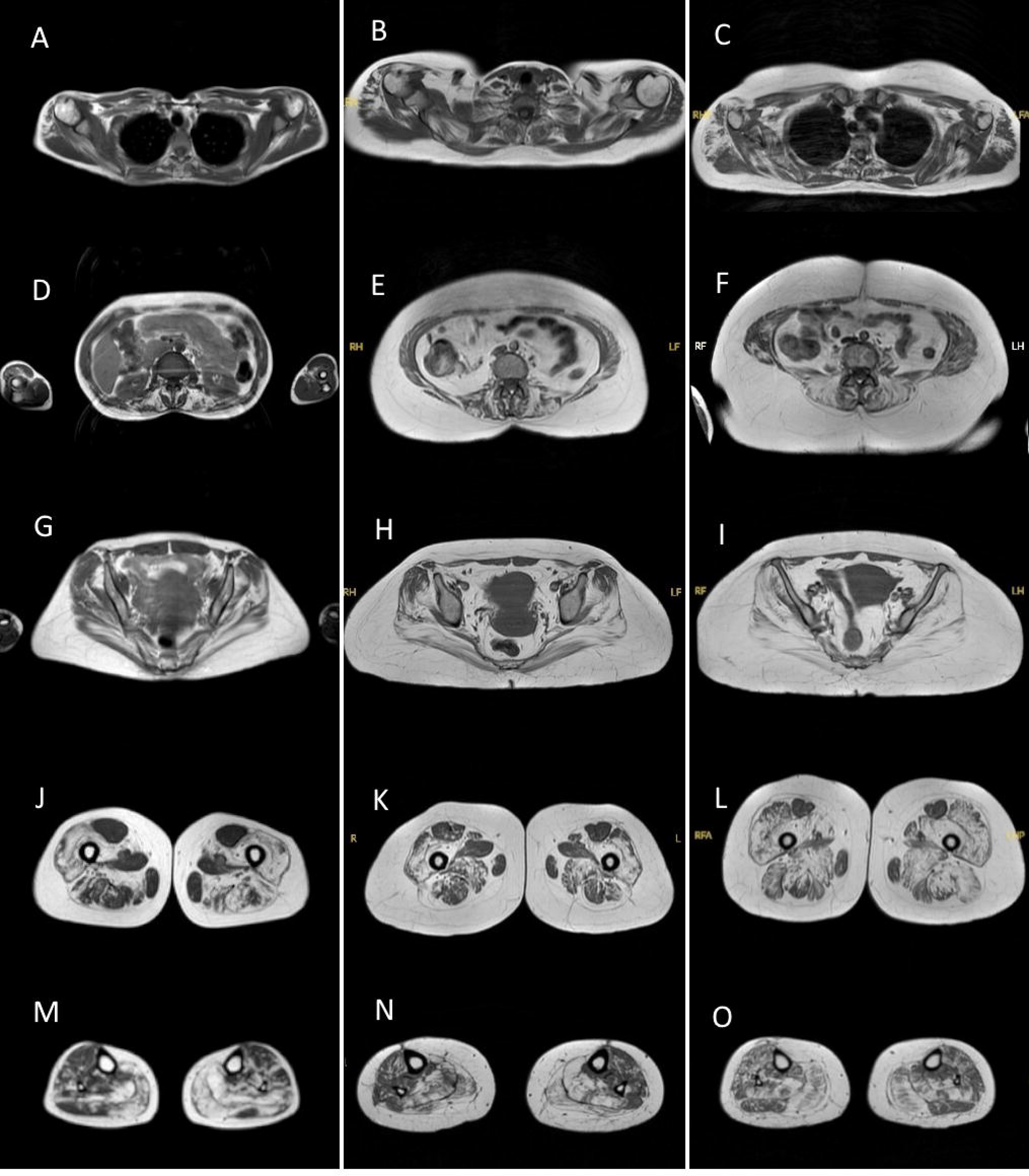
Whole-body muscle MRI from three patients. T1-weighted images from patients II:1 (proband), II:2 and II:3 (younger twins) are shown in each column revealing a similar pattern of muscle involvement. **(A-C)** Scapular girdle sequences show mild fat replacement mainly affecting deltoids, infraspinatus, and subscapularis muscles. **(D-F)** Thoracic and lumbar images show involvement of paravertebral muscles in all siblings. **(G-L)** Moderate to severe degree of atrophy and fat replacement in pelvic and thigh muscles, predominantly in glutei, quadriceps and posteromedial compartments. Note the remarkable sparing and hypertrophy of adductor longus and rectus femoris muscles, present in all cases. **(M-O)** At the leg level, calf muscles (soleus and gastrocnemii) are more affected than the anterolateral compartment. Finally, note the mild asymmetry of involvement present in some muscles (deltoids in A and B, glutei muscles in G and I, hamstring muscles in K and L, soleus in N, leg anterior compartment in O).

### Molecular genetics

No candidate variants were identified after sequencing a panel of 137 myopathy-causative genes, nor after exome sequencing. Genome sequencing identified a homozygous missense variant in exon 9 of *ATP2A2* (c.1117G>A, p.(Glu373Lys)) in the four affected siblings. Sanger sequencing showed the youngest unaffected individual and both parents were heterozygous carriers of the same variant. *ATP2A2* displays missense constraint (Z=7.02) and this variant is present once in gnomad v4 (allele frequency of 0.00000159).

### Protein modelling

Multiple sequence alignments were performed for SERCA2 orthologs across nine species (Fig.6A), anchored to the human SERCA2 protein sequence. The identified missense substitution affects a highly conserved amino acid residue, conserved till fruit fly. The “mutagenesis” function on PyMol was used to predict changes in SERCA2 protein structure and/or folding upon substituted amino acid residue incorporation into the sequence (Fig.6B). Both the wild-type glutamine and substituted lysine residues have side chains of similar length. However, the glutamine side chain is polar and uncharged, while the lysine residue is positively charged. Interestingly, amino acid substitution resulted in no changes to the polar contacts on either the side chain or protein backbone (Fig.6B).

**Figure 6.**
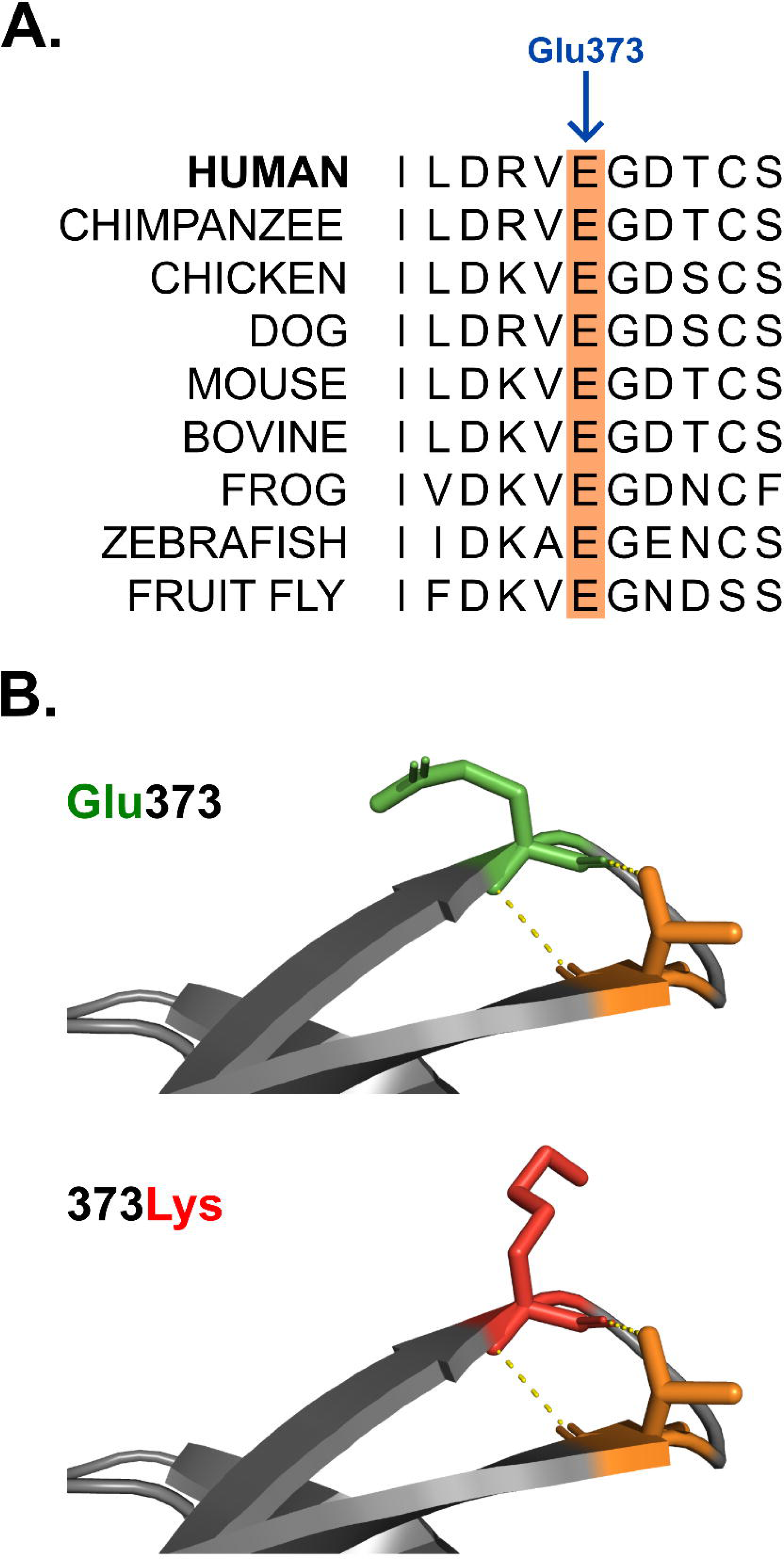
*In silico* investigation of the *ATP2A2* p.Glu373Lys variant. **(A)** Multiple sequence alignment of SERCA2 orthologs shows conservation of the identified variant in different species (E=glutamic acid). **(B)** The effect of the identified missense variant on three-dimensional SERCA2 structure. The wild-type residue is coloured green and mutant residue in red. Polar contacts with the Thr376 residue (orange) are shown with a yellow dotted line.

### Functional studies to assess Ca^2+^ kinetics after sarcolemma depolarization in myoblasts and myotubes

*In vitro* functional studies were performed in patient myoblasts and myotubes to assess kinetics of intracellular Ca^2+^ transport in response to electrical field stimulation or transient caffeine exposure. Myoblasts did not respond to field stimulation or caffeine exposure, neither from the patient nor from healthy controls (see supplementary figure S1). In myotubes, functional studies revealed that Ca^2+^ reuptake into the SR was slower in those from the patient compared to healthy controls. Thus, when subjected to electrical field stimulation every 20s, both the peak and baseline fluorescence was higher in myotubes from the patient compared to healthy controls. By contrast, the amplitude was smaller and the time to peak and the decay of the Ca^2+^ transient was slower in myotubes from the patient (Fig.7A). Moreover, when stimulated at increasingly shorter stimulation intervals, myotubes from the patient only responded to each stimulus with a stimulation interval of 20s. At shorter intervals, the slow Ca^2+^ removal from the cytosol resulted in a tetanic response of the Ca^2+^ signal in most myotubes from the patient. Wilcoxon’s rank-sum test showed that the number of tetanic responses at 20s, 10s, 3s and 1s was significantly higher in myotubes from the patient than from healthy controls (p=0.01). Fig.7B shows the average Ca^2+^ signals from 10 control myotubes and 13 patient myotubes stimulated at 20s, 10s, 3s and 1s intervals. Fig.7C summarizes the ability of myotubes to respond to the stimulation pulses at different pacing intervals and the fraction of myotubes exhibiting a tetanic response. Importantly, Ca^2+^ accumulation was not perturbed by the stimulation protocol as Ca^2+^ levels always returned to baseline when myotubes from the patient were left at rest before the next pacing interval (supplementary figure S2).

**Figure 7.**
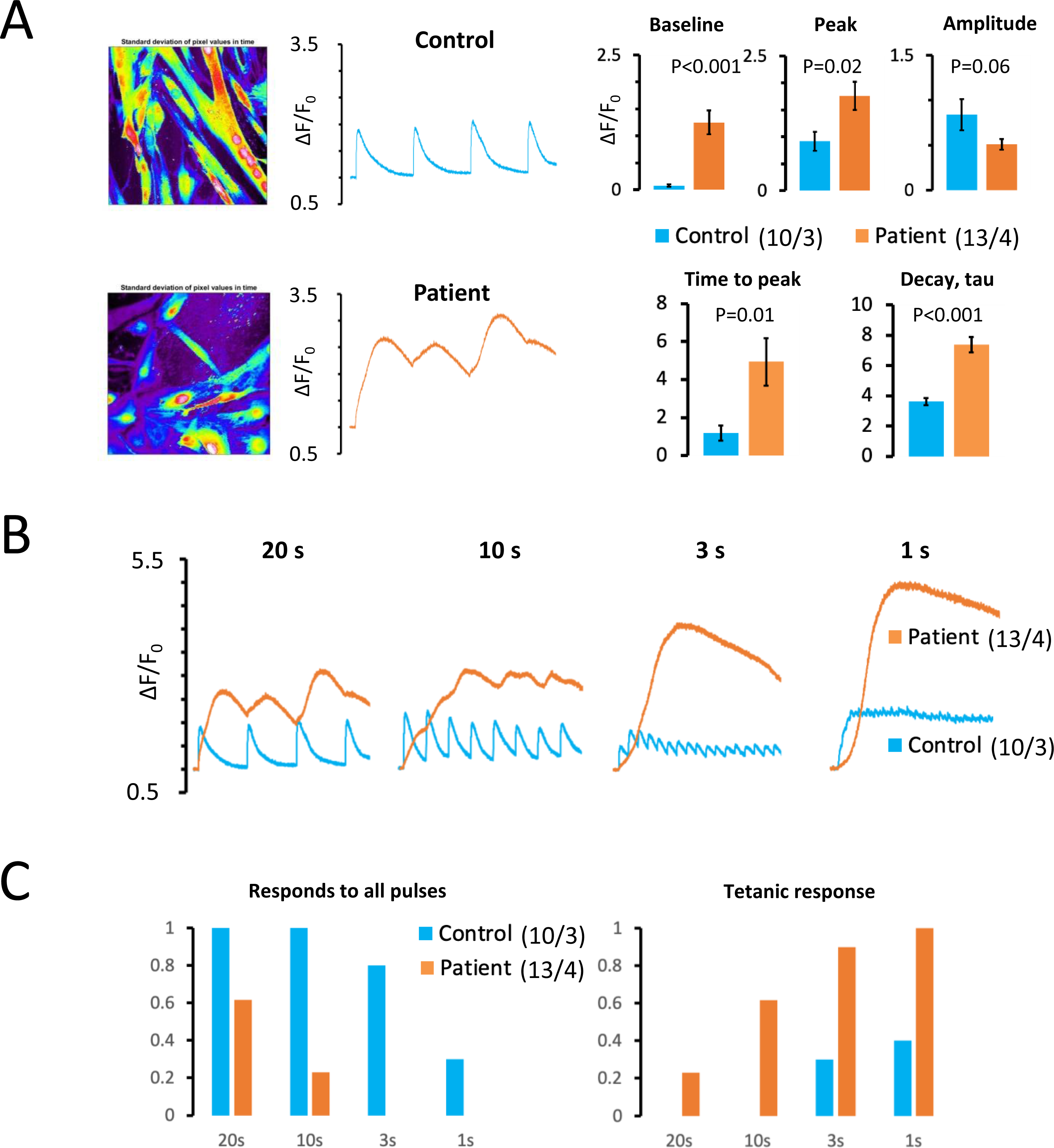
Functional studies showing intracellular Ca^2+^ transients in patient and control myotubes after electrical field stimulation. **(A)** Images of a field of myotubes from a control and the patient are shown on the left. Mean Ca^2+^ traces for 10 myotubes from healthy controls and 13 myotubes from the patient, subjected to electrical field stimulation at 20s intervals, are shown in the center and mean values for baseline Ca^2+^ and the peak, amplitude, time to peak and decay time constant (tau) of the Ca^2+^ transients are shown on the right. **(B)** Mean Ca^2+^ traces for myotubes from controls and the patient subjected to field stimulation at decreasing stimulation intervals. Numbers in parentheses indicate the number of myotubes and image fields analyzed respectively. **(C)** The fraction of myotubes that responded to all stimulation pulses are shown on the left for the different stimulation intervals (given below bars), and the fraction of myotubes showing a tetanic response is shown on the right.

To determine more directly the Ca^2+^ accumulating capacity of the SR in myotubes from the patient and from healthy controls, myotubes were exposed to two consecutive caffeine pulses (which releases the SR Ca^2+^ content into the cytosol) as outlined in Fig.8A. Analyzes of the amplitude and decay of the caffeine-induced Ca^2+^ transients (Fig.8B) revealed that the amplitude of the Ca^2+^ transient elicited by the first caffeine exposure was significantly larger in patient myotubes than in those from healthy controls, suggesting that myotubes from the patient have a greater Ca^2+^ storage capacity. By contrast, the decay of the Ca^2+^ transient during 10s after the first caffeine pulse was almost negligible in the patient while myotubes from healthy controls were able to reduce the Ca^2+^ transient by 49±8%, suggesting that the Ca^2+^ reuptake rate is strongly impaired in the patient myotubes. Consequently, the Ca^2+^ level after 10s was dramatically higher in the patient than in healthy controls, explaining why most myotubes from the patient displayed a tetanic response when stimulated every 10s, whereas myotubes from healthy controls did not produce a tetanic response to consecutive stimulation pulses (see Fig.7C). Moreover, the response of control myotubes to the caffeine pulses was consistent, while the response of myotubes from the patient was more heterogeneous with some displaying a very slow decay of the Ca^2+^ transient and lack of response to the second pulse while others showed a moderate decay of the first caffeine transient and a clear response to the second caffeine pulse (see supplementary figure S3), explaining why 20% of the myotubes responded to all pulses when stimulated every 10s and only 60% showed a clear tetanic response (see Fig.7C).

**Figure 8.**
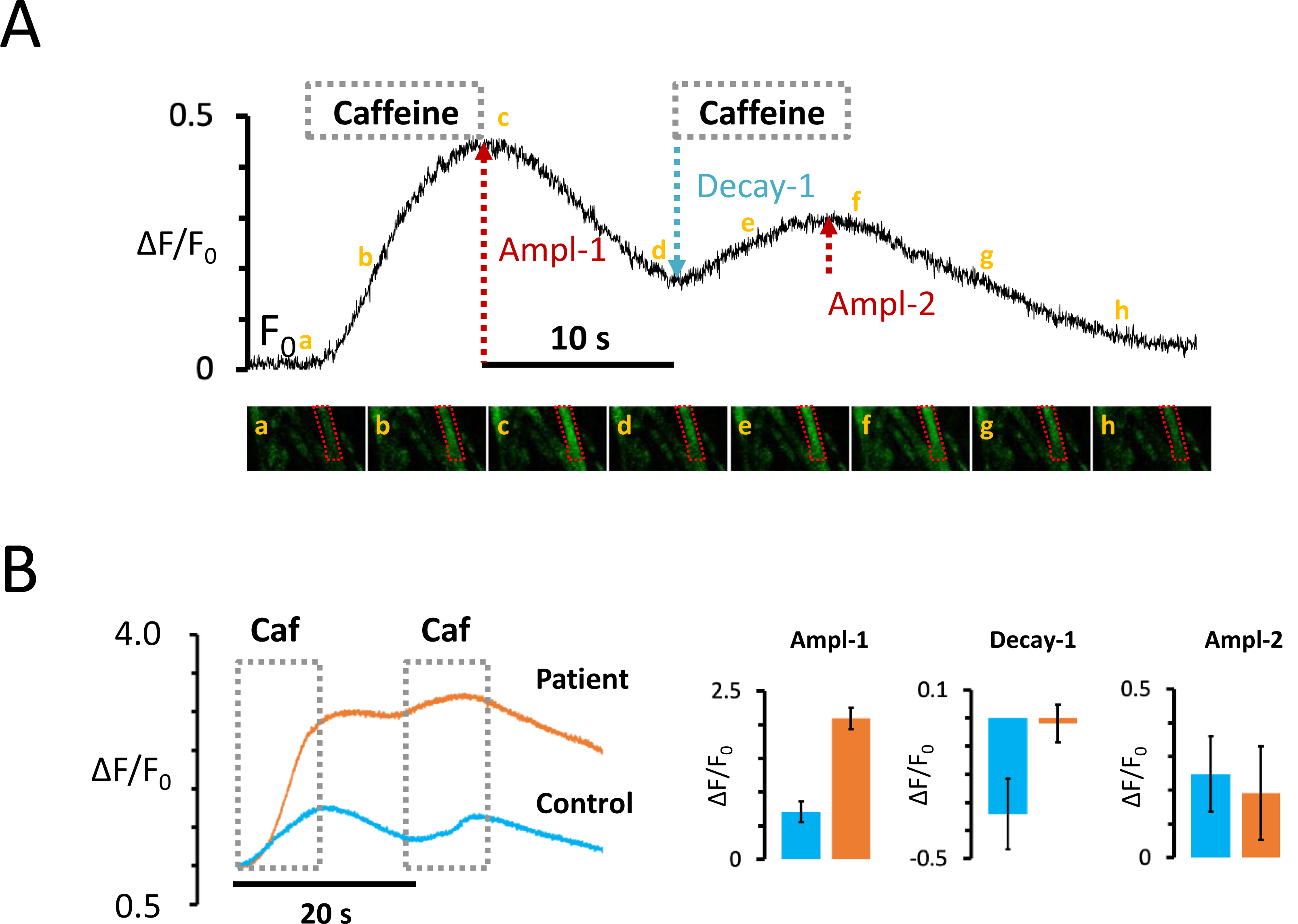
Functional studies showing intracellular Ca^2+^ transients in patient and control myotubes after caffeine exposure. **(A)** Representative Ca^2+^ recording from a control myotube subjected to two consecutive caffeine applications (indicated with gray boxes). The amplitude (Ampl-1) and decay (Decay-1) of the first caffeine induced Ca^2+^ transient and the amplitude (Ampl-2) of the second transient were measured as indicated. **(B)** Mean Ca^2+^ traces for 10 myotubes from healthy controls and 13 myotubes from the patient are shown on the left. The mean amplitude and decay of the first transient as well as the amplitude of the second transient are shown on the right.

## Discussion

The present study reports a family with four individuals affected with a novel autosomal recessive proximal myopathy caused by a variant in *ATP2A2*, the gene which encodes the Ca^2+^ channel SERCA2. The disease is clinically characterized by adult-onset proximal weakness affecting lower and upper limbs, moderately high CK levels, and absence of cardiac or skin involvement. Muscle biopsy revealed membrane-limited vacuoles only present in type I (slow-twitch) myofibers, where SERCA2a is enriched. Under electron microscopy, dilatations of the tubular system, proliferation of SR and some autophagic vacuoles were characteristically seen. Immunohistochemistry of SERCA2 suggested a redistribution of the protein in patient myofibers whereas WB revealed normal protein levels.

The Ca^2+^ handling *in vitro* studies performed in patient myotubes demonstrated an impaired function of SERCA2 resulting in altered Ca^2+^ homeostasis which is highly suggestive that the biallelic variant is pathogenic even though it does not have a protein destabilizing effect and is not located in known important functional or structural domains of the protein. Based on the functional studies, we presume that the *ATP2A2* substitution cause dysfunction of the SERCA2a pump in skeletal muscle and leads to the proximal vacuolar myopathy. Both patient and control myoblasts did not respond to field stimulation or caffeine exposure (see Results section), despite that SERCA2 is expressed in primary myoblasts and myotubes (unpublished data). The lack of response may be due to the fact that the excitation-contraction coupling complex is not sufficiently structurally developed in myoblasts(35). In developing myotubes, SERCA2 is much more abundant than SERCA1 (unpublished data), thus we can infer that the perturbed Ca^2+^ handling recorded in patient myotubes, compared to healthy controls, is due to the presence of mutant SERCA2 pumps.

This novel myopathy should be included in the group of disorders of excitation-contraction coupling and Ca^2+^ homeostasis abnormalities, to which the majority of congenital myopathies belongs(36,37). SERCA2 is critical for muscle relaxation to pump Ca^2+^ back into the SR after closure of RYR1 channels. Our *in vitro* studies prove that cultured patient myotubes have an increase in cytosolic Ca^2+^ levels after repeated contractions, as happens in the gain-of-function *RYR1* mutations leading to “leaky channels”, which are generally linked to central core disease(38,39). Despite this similarity in resting Ca^2+^ concentrations, muscle pathology and phenotype are clearly different in SERCA2-related myopathy compared to RYR1-related disease.

Interestingly, despite SERCA2a expression in cardiac tissue, our patients did not present cardiomyopathy. The fact of absent objectifiable cardiomyopathy to date is surprising and should be explored in families with the same condition. A possibility, given that cardiac muscle expresses a higher amount of SERCA2a than slow-twitch skeletal myofibers(5), is that the pump functional impairment proven in our studies may not affect sufficiently Ca^2+^ homeostasis in cardiomyocytes to cause cardiac dysfunction. Likewise, neither of the siblings had skin or nail abnormalities that could relate the myopathy with DD or represent a phenotypic overlap between DD and proximal myopathy. The clinical phenotype of pure skeletal myopathy and the findings reported here suggest the possibility that c.1117G>A variant only (or mainly) affects the synthesis and/or function of the isoform SERCA2a which is expressed at higher levels in type I skeletal myofibers, while SERCA2b is widely expressed in other tissues including the skin. However, in our opinion, no genetic explanation can as yet support this hypothesis as the missense variant c.1117G>A is localized in exon 9 and the only difference in *ATP2A2* transcripts a and b is the 20^th^ exon. A larger exon 20 leads to an extra 11^th^ transmembrane domain in isoform 2b that forms the 2b-tail(40). Another explanation of the absent skin involvement in our patients could be the physiologically slower Ca^2+^ transport rate of SERCA2b attributable to the 2b-tail, which seems to be beneficial for keratinocytes(15,41). In fact, a recent study of a Darier-causing variant proved that skin mutant cells exhibit lower Ca^2+^ affinity and a higher transport rate leading to skin disease(42); thus, it could be stated that the variant found in our study causes a decrease in the activity of both SERCA2 isoforms, but results in pathology only in skeletal myofibers, without pathological consequences in keratinocytes. Either way, the absence of cardiomyopathy should be further explored as cardiac muscle expresses the same SERCA pump.

Of the 270 variants in *ATP2A2* associated with DD, there are many missense variants in addition to essential splice site variants, frameshift and nonsense variants. This suggests that in DD a likely mechanism may be haploinsufficiency. *ATP2A2* is under loss-of-function constraint (pLI =1, gnomAD) supporting the data from DD that haploinsufficiency is likely to be associated with disease. Given that SERCA2 protein abundance is not reduced in our patient skeletal muscle, this suggests that the mutant protein produced from the variant is likely behaving as a functional hypomorph, or via a toxic gain of function.

Regarding the muscle pathology features, sarcotubular myopathy resulting from variants in *TRIM32* (43–46) should be included in the differential diagnosis of the disease here described, due to its similar histological findings in mice and human muscle, including autophagic vacuoles and dilated sarcotubular structures. Variants in *TRIM32*, which encodes the Tripartite Motif-Containing Protein 32, cause an autosomal recessive muscular dystrophy called limb-girdle muscular dystrophy R8 or sarcotubular myopathy (LMGDR8/STM). TRIM32 is a multifunctional protein with an E3 ubiquitin ligase activity that regulates degradation of many target proteins(47). In mouse primary skeletal myotubes, TRIM32 participates in cellular Ca^2+^ transport by a ubiquitin-independent binding to SERCA1 via its NHL repeats, which enhances SERCA activity and increases Ca^2+^ level in the SR(48). This is a very interesting finding because it could link physiopathological mechanisms triggering vacuole formation both in TRIM32 deficiency and SERCA2a functional deficiency. Also, it is possible that SERCA pumps and TRIM32 are correlated with each other via notch signaling in skeletal muscle(49–51). Clinically comparable to the patients described in this study, patients with *TRIM32* variants normally develop progressive weakness of the lower limbs with no cardiac or respiratory symptoms.

### Conclusions

We report a novel adult-onset autosomal recessive vacuolar myopathy caused by a homozygous variant in *ATP2A2*, characterized by limb-girdle weakness. Biopsy findings show presence of vacuoles restricted to type I myofibers, corresponding to dilatation and proliferation of the T-tubule system. Given that SERCA2 is abundant in type I myofibers and the muscle pathology is highly restrictive to these same myofibers, the variant found in *ATP2A2* is likely causative of the muscle disease in this family. Assessment of Ca^2+^ dynamics *in vitro* in cultured control and patient myotubes showed a clear reduction of the SERCA2 pumping capacity and higher resting Ca^2+^ levels in the patient myotubes, further supporting the pathogenicity of the biallelic c.1117G>A variant leading to an altered excitation-contraction coupling in skeletal muscle.

It remains to be seen whether other variants in *ATP2A2* also lead to a myopathic phenotype. We recommend this gene be included in genomic investigations in patients with myopathy, especially when the muscle pathology shows similar features to those described here. The identification of additional patients with SERCA2-related myopathy will clarify the genotype-phenotype correlations associated with *ATP2A2* variants.

## Data Availability

All data produced in the present study are available upon reasonable request to the corresponding author.
Data including exact ages, gender or place of origin has been removed following Medrxiv guidelines. This fact includes removal of Figure 1 (family pedigree) from this version to avoid possible identification of the patients. All this information can be available upon request.

## Declarations

### Ethics approval

not applicable.

### Consent for publication

Written consent to perform muscle biopsy and NGS studies was taken from the corresponding individuals, as well as consent for publication from the affected individuals.

### Availability of data and material

The data supporting the findings of this study are available from the corresponding author upon request.

## Authors’ contributions

Conceptualization: MO, GR, EG, NL; Methodology: MO, GR, CA, JP, EG, LH, LL, RC; Investigation: MO, GR, LH, CA, AG, EG, LL, CA, PG, JP, RC, MC, AC, AV; Funding acquisition: MO, GR; Supervision: MO, GR, NL, EG; Writing– original draft: LL, GR, LH, MO; Writing– review and editing: MO, GR, NL, LH, EG, LL. All authors read and approved the manuscript.

## Acknowledgements

We are extremely grateful to all the family for the collaboration in this project. We further thank the electron microscopy core facility in Universitat de Barcelona.

