## Supplementary table for "A homozygous *ATP2A2* variant alters sarcoendoplasmic reticulum Ca^2+^-ATPase 2 function in skeletal muscle and causes a novel vacuolar myopathy"

**Supplementary Table 1.** Antibodies used in the present study.

| **Antigen/ Reference** | **Epitope** | **Host** | **Dilution (IH/IF)** | **Dilution (WB)** | **Manufacturer** |
| --- | --- | --- | --- | --- | --- |
| SERCA2/  MA3-919 | Monoclonal | Mouse | 1:100 | 1:1000 | Invitrogen |
| SERCA1/  MA3-912 | Monoclonal | Mouse | 1:100 |  | Invitrogen |
| RYR1/  R129 | Monoclonal | Mouse | 1:1000 |  | Sigma-Aldrich |
| P62/  BML-PW9860-0100 | Polyclonal | Rabbit | 1:200 |  | Enzo Life Sciences |
| Dystrophin Carboxi/  DYS 2 | Monoclonal | Mouse | 1:100 |  | Novocastra |
| Dystrophin Rod/  DYS 1 | Monoclonal | Mouse | 1:2 |  | Novocastra |
| Caveolin-3/  610421 | Monoclonal | Mouse | 1:20 |  | BD Transduction |
| Dysferlin/ NCL-Hamlet | Monoclonal | Mouse | 1:100 |  | Novocastra |

**IH** immunohistochemistry; **IF** immunofluorescence; **WB** western blot.

**Supplementary Table 2.** UniProt accession codes used for Multiple Sequence Alignment of SERCA2 orthologs.

| **Protein Sequence** | **Corresponding gene name** | **Animal species** | **Accession code** | **Uniprot review status** |
| --- | --- | --- | --- | --- |
| HUMAN | *ATP2A2* | *Homo sapiens* | P16615 | Reviewed |
| CHIMPANZEE | *ATP2A2* | *Pan troglodytes* | K7BZQ7 | Unreviewed |
| MOUSE | *Atp2a2* | *Mus musculus* | O55143 | Reviewed |
| BOVINE | *ATP2A2* | *Bos taurus* | F1MPR3 | Unreviewed |
| DOG | *ATP2A2* | *Canis lupus familiaris* | O46674 | Reviewed |
| CHICKEN | *ATP2A2* | *Gallus gallus* | Q03669 | Reviewed |
| FROG | atp2a2 | *Xenopus tropicalis* | A0A6I8SKA5 | Unreviewed |
| ZEBRAFISH | atp2a2b | *Danio rerio* | Q6ZM60 | Unreviewed |
| FRUIT FLY | *SERCA* | *Drosophila melanogaster* | P22700 | Reviewed |
