## supplementary figure for "A homozygous *ATP2A2* variant alters sarcoendoplasmic reticulum Ca^2+^-ATPase 2 function in skeletal muscle and causes a novel vacuolar myopathy"

### Supplementary figures S1-S3

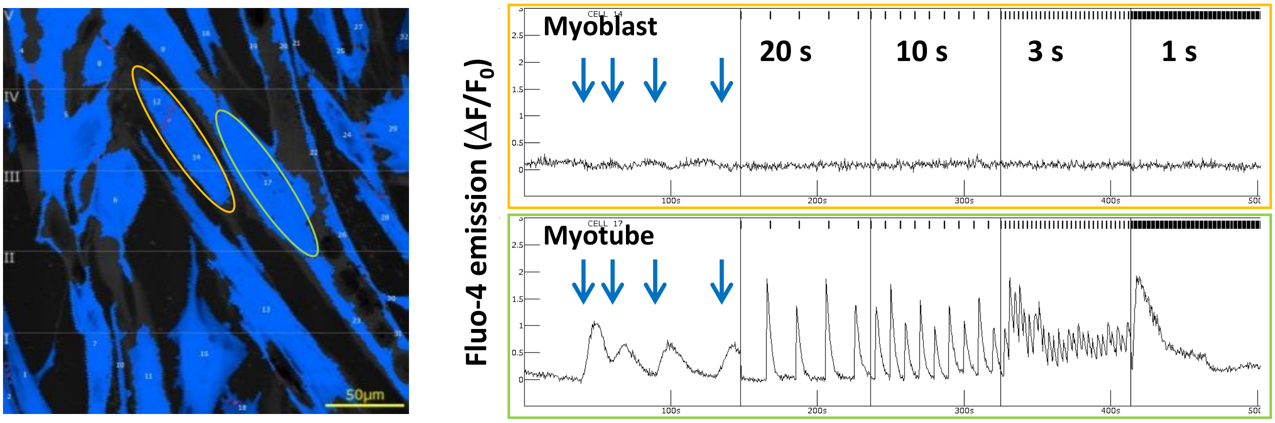


**Figure S1.** Representative calcium recordings from a myoblast (orange label) showing no response to caffeine (blue arrows) or electrical field stimulation; and a myotube (green label) showing a clear response to caffeine and field stimulation.

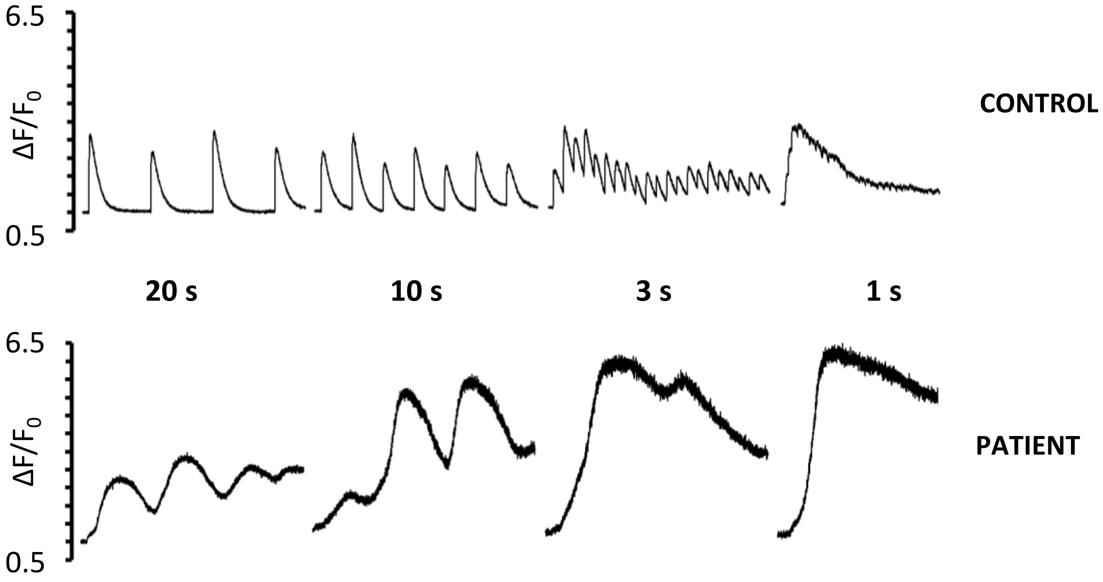


**Figure S2.** Calcium recordings from a control myotube (top) and a patient myotube subjected to field stimulation at 20s, 10s, 3s and 1s stimulation intervals. Myotubes were left at rest between each stimulation frequency to allow calcium levels to return to baseline before the next stimulation frequency.

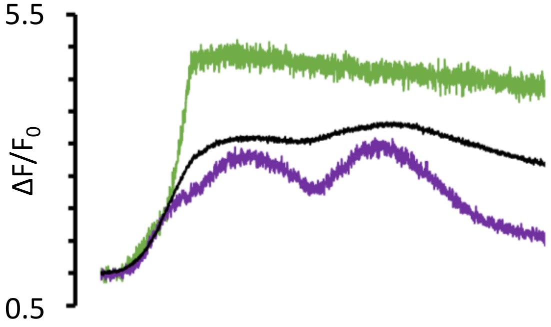


**Figure S3.** Representative calcium recordings from myotubes from the patient, subjected to two consecutive caffeine applications. Green and purple traces illustrates differences among myotubes and the black trace represents the average response of 13 myotubes from the patient.
